# Priority Setting of Ventilators in the COVID-19 Pandemic from the public’s perspective

**DOI:** 10.1101/2020.06.10.20127290

**Authors:** Fariba Asghari, Alireza Parsapour, Ehsan Shamsi Gooshki

## Abstract

**Background:** Ventilator allocation plan for public health crisis should be developed through recognizing the values of society and engaging the general public. This study was conducted to assess the Iranian citizens’ attitude about a number of principles and criteria for allocation of ventilators in current COVID-19 epidemic.

**Materials and Methods:** An electronic self-administered questionnaire was publicly distributed through social networks of Telegram and WhatsApp to perform this cross-sectional study. The questionnaire consisted of 11 statements about the selection and prioritization of patients for the use of a ventilator.

**Results:** 1262 persons, including 767 citizens and 495 health care providers participated in this study. More than 95% of participants agreed upon the necessity to avoid discrimination and avoid prioritization according to patients’ gender, economic and political status. While 40.9% of citizens and 49.6% of healthcare workers believed that a ventilator can be disconnected from a patient with a poor prognosis to help a patient who has a better prognosis (P-value=0.13), 34.3% of people and 29.6% of healthcare workers believed that the earlier admitted patients have the right to receive the device even if the likeliness of his/her survival is less than the next patient (P-value=0.009).

**Conclusions:** This study showed that people accept maximizing health benefits as a measure of ventilator allocation in the pandemic of COVID-19. At the same time, periodic evaluation of patients and disconnecting the device from a patient that no longer benefits from ICU services requires its scientific and ethical basis to be brought in public discourse.

## Introduction

The COVID-19 epidemic raised concerns about the lack of adequate health resources in most countries. One of the most difficult decisions in this regard is to allocate life-sustaining treatments mainly ventilators and intensive care units for the surge of patients with respiratory failure. Generally, in public health crisis, when healthcare resources for all patients are short, the basic principle of providing best care for the patient shifts to maximizing the benefit to the whole affected population(1), so that the maximum number of lives could be saved. No single criterion can cover all relevant ethical values for triage, and a combination of different criteria should be considered in the priority setting of resource to meet healthcare needs (2). The general principles in resource allocation are fair access and utility maximization (3). However, in interpreting these two principles, other criteria such as age, saving more quality-adjusted life years (long-term survival), prioritizing people who are needed for maintaining the function of society in public health crises are discussed, all of which are serious issues (4).

Policy-making for resource allocation must be done by the participation and involvement of stakeholders during epidemics. Paying attention to the general public and engaging them in decisions is essential for establishing trust and maintaining social order (5, 6). Developing resource allocation policies should be done before the occurrence of public health crisis and through engaging the stakeholders and the general public (7, 8). The lack of preparedness and hasty policymaking in the allocation of intensive care due to the COVID-19 crisis has risen concerns which have even led to several protests by social activists in Italy and several US states about unfair resource allocation for a group of patients (9-14). Lack of transparency and non-involvement of the society in the policy-making process of planning for scarce resource allocation could raise public concerns about the fairness of providing care to patients (15).

Fortunately, in Iran, we have not reached this level of limitation in intensive care services and ventilators. However, we, unfortunately, have no policy on the allocation of intensive care in the surge of epidemics in Iran. In an epidemic crisis, one of the policymakers’ concerns is avoiding panic in the society so they might prefer not to encourage public debate about such sensitive issues. Because of such concerns the epidemic strike of COVID-19 might not be considered to be the right time to engage the community in patient triage policy for receiving ventilator. However, on the other hand, policies on scarce recourses allocation cannot be developed without obtaining public concerns and opinions. To have a better understanding of the public point of view and also provoke discussion about this issue, we decided to obtain the people’s viewpoints on the different criteria for allocating ventilators in the current epidemic crisis. Nonetheless, public polls are not a good way for public engagement in resource allocation decisions, it can be considered as a starting point for recognizing public sensitivities, concerns, and misconceptions about priority setting that may help policy-makers. As, public surveys have been conducted in some public engagement in healthcare policy (16).

## Materials and methods

### Research design and tool

This cross-sectional study was conducted using a self-administered questionnaire with distribution via social networks of WhatsApp and Telegram. The study questionnaire was developed by the authors based on an overview of the current challenges in ventilator allocation in the pandemic of COVID-19. Primary questions were designed as 19 statements, that each one reflected a criterion for selecting and prioritizing patients in receiving a ventilator so that participants could express their agreement or disagreement about each statement. Questions were sent to 10 citizens who were not healthcare workers (convenience sampling) to assess the transparency and comprehensibility, and they were asked to rate each of the statements in terms of transparency and comprehensibility.

**Box 1. Ventilator allocation statements**

1. Patients who have more likeliness of survival should prioritized to be benefited from to the device. (survival)
2. Until all ventilators are occupied, each patient who needs mechanical ventilation should be connected to a ventilator. (Duty to care)
3. With equal chance of successful treatment, the patient’s quality of life should not be the basis of letting to use the ventilation device. It means that severely mentally or physically disabled people should have equal rights with ordinary people. (quality of life)
4. When all devices are occupied and a new patient who is more likely to survive needs ventilation, the device should be disconnected from the patient who is less likely to survive and connected to the new patient. (reassessment of patient)
5. The patients who has earlier need for the device has the right to use it, even if he/she is less likely to survive than the next patient. (first come first served)
6. A patient who has no family or relatives or no one is asking about him/her should not be deprived of receiving the device only because of this reason. (social support)
7. The economic status of patients should not be considered as a basis for selecting their priority for being able to use the device. (financial ability)
8. The patient’s political position (for example, being a member of parliament) should not be considered a criterion for being able to use the device. (political position)
9. It is fair that those who serve patients of Corona virus, such as physicians and nurses should be given priority for using the device if become sick. (reciprocity)
10. There should be no difference between men and women if the chance of success of treatments are the same. (gender equity)
11. If the severity of the disease and the probability of successful treatment are the same between two patients who are of different ages, for example, one is 30 years old and the other is 50 years old, or one is 50 years old and the other is 70 years old, and there is only one device available, priority should be given to the person with the lower age. (Younger patient)

To evaluate the content validity, questionnaire statements were sent to 20 experts in the field of medical ethics, emergency medicine, and ICU care, 16 of whom responded and assessed the validity of each statement as a criterion for allocation of ventilators. Based on the experts’ answers, the content validity ratio (CVR) index was calculated for each of the statements, and those statements that their CVR were less than critical value based on the Lawshe table omitted from the questionnaire. Besides that, the experts were asked to state their comments to improve the narration of each statement. The revised statements were sent back to experts for CVR assessment. Eventually, after omitting 8 questions, 11 statements remained in the general survey on the criteria for ventilator allocation in the Coronavirus crisis (Box 1).

The final statements were again sent to 6 people outside the health care system to evaluate the transparency and comprehensibility, all of whom stated that the statements were understandable.

In addition to these statements, a number of questions were added to the final questionnaire about demographic characteristics, place of residence, and history of infection or hospitalization of oneself or first-degree family members because of coronavirus, as well as employment as health care provider. The questionnaire was designed electronically on the epoll^1^ platform.

### Patient and Public Involvement

Regarding ethical consideration, there was a concern that in the current event of the infectious epidemic, the questions of this study raised concerns among people about the lack of resources or the existence of unfair allocation of ventilators in Iran hospitals. In order to prevent this concern and avoid forming a media hype in this regard, we contacted people participating in face validity phase and asked them about their perceptions and feelings about the questions. Questions that would mislead the participants and imply that such problems really exist in hospitals and therefore raise concerns about unfair procedures in the allocation of health services were removed or changed from negative statement to positive one to avoid the emergence of concerns about unfair procedures in the allocation of resources in the health care system. As mentioned before, the COVOD-19 outbreak in Iran has not reached such level that obligate health care facilities to select among patients in need of ventilators. The second sample of participants who confirmed the transparency of the final statements stated that the nature of the questions do not imply such concerns for them. The pilot study was initiated by sending the questionnaire to 30 hospitalized patients of the coronavirus after being treated and discharged from the hospital. They were able to contact the researcher to ask or state anything about the survey. Distribution of the questionnaire started when we get ensured of not creating public panic or fear. The study protocol was approved by the research ethics committee of Tehran University of Medical Sciences under license ID IR.TUMS.VCR.REC.1399.085. Participation in the study was voluntary and anonymous, and this was mentioned in the announcement of the study.

The questionnaire was sent to WhatsApp and Telegram groups and channels -which are the main social media used by Iranian-to a large extent. The invitation letter asked the participants to introduce this survey to their friends in other groups and channels on social networks. The report of this poll was broadcasted in the ISNA news agency, which is one of the most popular news agencies in Iran(17). The study was completed after two weeks, with a decrease in the number of participants.

### Analysis

Although the main objective of the study was to examine the public opinion, due to the availability of medical social network groups and the high level of participation of health care providers, it was decided at the time of administration that in addition to the opinion of the general public, the difference between public opinion (Non health-care-worker citizens which afterwards will called citizens) and health care workers (HCWs) be compared. The opinions of the HCWs and the public were compared with the chi-square for trend.

Multiple regression analysis was used to evaluate the underlying variables for agreement or disagreement with each of statements. For this purpose, the dependent variables (participants’ opinion about each statement) were converted to two value of agreement and no agreement (disagreement or having no idea). To analyze the effect of the living place variable, the classification of provinces based on the index of information and communication technology development (access, use, and information technology skills) was used. Based on this index, the Information Technology Organization of the Ministry of Communications of Iran has divided the provinces into three categories. This classification was used to evaluate the influence of geographical location on the opinion of the participants (18). The participants’ distribution in the areas of information technology development was compared with the whole country population distribution in these areas.

## Results

A total of 1262 participants including 767 citizens (60.8%) and 495 HCWs (39.2%) from all provinces of the country, from 97 cities (citizens from 77 cities and HCWs from 73 cities) were enrolled in this study. The demographic characteristics of the two groups of health care providers and citizens are presented in Table 1. The two groups were different in terms of education, being infected with the coronavirus, and hospitalization due to the coronavirus. There was no significant difference in population distribution in terms of information technology development groups between citizens and health care providers, however, there was a significant difference between the two groups of participants in the study and the country’s general population (0.001). The average age (± standard deviation) of HCWs and the citizens’ population was 39.66 (± 12.36) and 40.63 (± 10.53) years, respectively, which was not significantly different.

**Table 1.** Demographic characteristics in health care workers and citizens

| characteristics |  |  | Citizens | HCWs | P-value |
| --- | --- | --- | --- | --- | --- |
| Gender | female | n | 477 | 282 | 0.064 |
|  |  | % | 62.4 | 57.2 |  |
|  | male | n | 287 | 211 |  |
|  |  | % | 37.6 | 42.8 |  |
| Education | High school or less | n | 11 | 0 | <0.001 |
|  |  | % | 1.4 | 0.0 |  |
|  | High school graduation/ college | n | 129 | 22 |  |
|  |  | % | 16.9 | 4.5 |  |
|  | BS | n | 281 | 57 |  |
|  |  | % | 36.7 | 11.5 |  |
|  | MS | n | 236 | 40 |  |
|  |  | % | 30.8 | 8.1 |  |
|  | PhD | n | 108 | 375 |  |
|  |  | % | 14.1 | 75.9 |  |
| History of COVID-19 infection | NO | n | 741 | 444 | <0.001 |
|  |  | % | 97.4 | 90.2 |  |
|  | YES | n | 20 | 48 |  |
|  |  | % | 2.6 | 9.8 |  |
| History of COVID-19 infection in a close family member | NO | n | 678 | 403 | <0.001 |
|  |  | % | 89.3 | 81.9 |  |
|  | YES | n | 81 | 89 |  |
|  |  | % | 10.7 | 18.1 |  |
| ICT Development Index (IDI) | 1 | n | 453 | 325 | 0.052 |
|  |  | % | 59.9 | 66.3 |  |
|  | 2 | n | 203 | 108 |  |
|  |  | % | 26.9 | 22.0 |  |
|  | 3 | n | 100 | 57 |  |
|  |  | % | 13.2 | 11.6 |  |

The most important agreed criteria in the allocation of ventilators from the viewpoint of people were avoiding prioritization based on gender, financial ability, and the political position of individuals, and more than 95% of people agreed on each of these views (Table 2). There was no agreement of more than 50% on prioritization based on the quality of life, first come first served, and allocation to younger patients. In other words, the participants had a serious difference of opinions on these values.

**Table 2.** Frequency and relative frequency of agreement with ventilator allocation criteria in the COVID-19 crisis in terms of the opinions of citizens and HCWs.

| criteria |  | citizens | HCWs | P-Value |
| --- | --- | --- | --- | --- |
| survival | n | 547 | 407 | 0.01 |
|  | % | 72.1 | 82.4 |  |
| Duty to care | n | 711 | 436 | <0.001 |
|  | % | 93.1 | 88.1 |  |
| Quality of life | n | 482 | 222 | <0.001 |
|  | % | 62.9 | 44.8 |  |
| patient reassessment | n | 312 | 245 | 0.13 |
|  | % | 40.9 | 49.6 |  |
| First come first served | n | 260 | 146 | 0.009 |
|  | % | 34.3 | 29.6 |  |
| Social support | n | 653 | 434 | 0.29 |
|  | % | 85.2 | 87.7 |  |
| Financial ability | n | 745 | 488 | 0.16 |
|  | % | 97.5 | 98.6 |  |
| Political position | n | 742 | 489 | 0.36 |
|  | % | 96.7 | 98.8 |  |
| reciprocity | n | 575 | 336 | 0.001 |
|  | % | 75.2 | 67.9 |  |
| Gender equity | n | 747 | 488 | 0.34 |
|  | % | 97.6 | 98.8 |  |
| Younger patient | n | 277 | 200 | 0.31 |
|  | % | 36.3 | 40.4 |  |

From the viewpoint of the majority of citizens and HCWs, the decision to allocate a ventilator should be made collectively by a group of physicians (n = 596) (78.1%) and (n = 430) 86.9%, respectively (p-value <0.001).

There was no significant relationship between the living area of the participants in terms of information technology development index and people’s opinions on the criteria for ventilator allocation.

The multiple analysis of regression to determine the factors affecting agreement with the criteria of economic ability and gender equality showed that none of the underlying variables have an effect on the agreement with these two criteria. Factors affecting other ventilator allocation criteria that remained in the regression equation are presented in Tables 3 and 4.

**Table 3.** Multiple regression analysis of ventilator allocation criteria with background variables

| criteria | factor | OR | 95% CI |
| --- | --- | --- | --- |
| Survival | HCW | 1.87 | 1.41-2.49 |
| Duty to care | Age | 1.04 | 1.02-1.05 |
|  | HCW | 0.53 | 0.36-0.79 |
|  | A History of covid-19 infection | 12.22 | 1.25-119.48 |
|  | History of hospitalization because of covid-19 | 0.046 | 0.004-0.51 |
| Quality of life | HCW | 0.51 | 0.41-0.65 |
| patient reassessment | HCW | 1.40 | 1.11-1.77 |
|  | Gender | 1.40 | 1.11-1.77 |
| First come First served | age | 1.01 | 1.00-1.02 |
| Social support | History of covid-19 infection in first-degree family members | 2.04 | 1.13-3.69 |
| Political position | HCW | 3.17 | 1.24-8.08 |
|  | History of covid-19 infection | 0.25 | 0.08-0.77 |
| reciprocity | Age | 1.01 | 1.00-1.02 |
|  | HCW | 0.64 | 0.50-0.83 |
|  | History of covid-19 infection | 2.40 | 1.15-5.02 |
|  | History of hospitalization because of covid-19 | 0.25 | 0.07-0.89 |
| Younger patient | education | 0.54 | 0.36-0.80 |
|  |  | 0.64 | 0.45-0.91 |

**Table 4.**
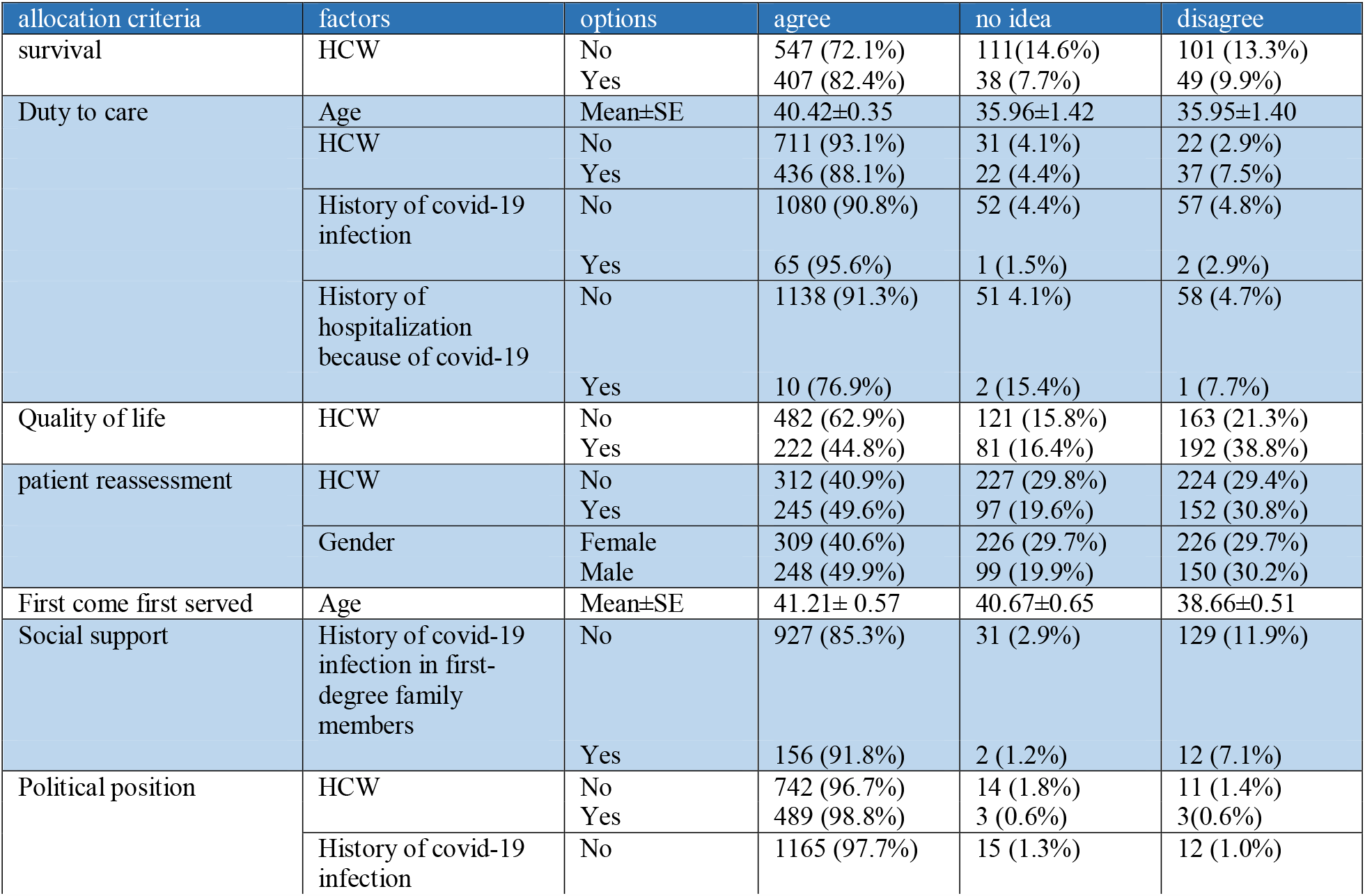
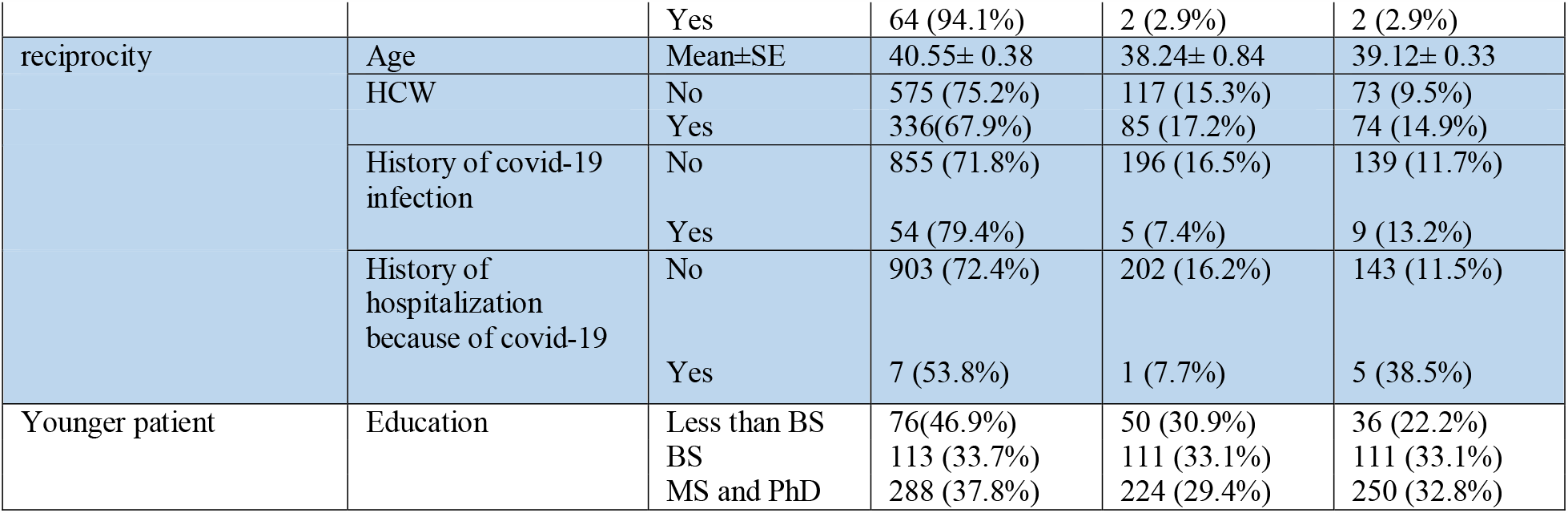
The level of agreement with the allocation of ventilators based on the factors entered in the regression equation

Still, being a health care worker was the most important determining factor in participants’ attitudes toward allocation criteria. Participants’ level of education was only effective in prioritizing of younger patients, however, this effect was not linear, so that until the bachelor’s degree, the dominating belief was to prioritize younger patients, in contrast, with the increase in the education level, the opinions about prioritization was inverse. Gender was only an influencing factor in the disconnection of the ventilator from a person with a low prognosis to save a person with a better prognosis (Table 4). The most predicting effect was the hospitalization factor due to corona infection on the criteria of non-prioritization until the capacity of beds for patients who need it, as well as reciprocity value, which showed that hospitalization due to corona infection reduces agreement with these criteria (Table 4).

## Discussion

The objective of this study was to evaluate the public opinion regarding the various criteria that can be considered in the allocation of ventilators. This study was conducted for the first time in Iran, and based on the review of the literature, no similar study was found in other countries that specifically evaluated the viewpoint of people on ventilator allocation during the COVID-19 epidemic crisis, so there was no data available for comparison and benchmarking.

The results of this study showed that the viewpoint of citizens toward ventilator allocation criteria during the COVID-19 crisis is generally consistent with the viewpoint of HCWs. The most common consensus of the citizens was that the financial and political position, as well as gender, should not be used as a basis to prioritize patients. Unfortunately, at the beginning of the COVID-19 epidemic, despite the limited resources of protection equipment, Iranians received news about out-of-protocol access to protective equipment and PCR testing for politicians and government officials (19). This raised concerns about unfair access to services. The findings of this study showed that the patients of COVID-19 themselves were more likely to disagree with this criterion.

Except for the three criteria of prioritizing the younger, comes first served first, and saving the patient with better prognosis, which had a serious controversy upon, there was an agreement of more than 50% on other statements of this questionnaire. Participants of this study disagreed on prioritizing COVID-19 patients based on the quality of life, while a study showed that 74% of people in England agreed that quality of life should be considered in providing life-saving care (20). However, that study was conducted at a very different time from our study. It seems that the context of the infectious disease epidemic, which is not related to the previous patient’s quality of life, has led to the difference on this issue in the present study.

For the allocation of the ventilator to a person with a better prognosis, citizens, like HCWs, had a consensus on this criterion, although this belief was more robust among HCWs. In general, working in the health care profession has an effect on the opinion of saving a patient with better prognosis, prioritizing based on better quality of life, disconnecting the device for saving a patient with better prognosis. It seems that in order to make their work meaningful, HCWs are more inclined to save the lives of patients with more life expectancy.

93% of citizens agree that as long as all ventilators are not occupied, every needy patient should be connected to the device, which shows that citizens do not want these policies to be implemented early and it is probably an expression of the need for regional, instead of hospital policies for ventilator allocation(1). Since the lack of information about the policies of other hospitals in the region and the condition of their bed occupations may cause early priority policy initiation rather than patient transfer to other hospitals (21).

Although the prioritization of health resources based on social support has been mentioned as a criterion in organ allocation (22, 23), this study showed that this criterion does not apply for the need of patients for ICU services in epidemics of acute respiratory disease. In the opinion of the participants, a patient who has no family or relatives should not be deprived of receiving the device only for this reason.

There was also consensus on the criterion of reciprocity, or prioritization of patients such as physicians and nurses who serve to help patients of the coronavirus as their gratitude. This might not be the case about whoever involved in epidemic control. It is interesting to note that were more likely to agree to this criterion than HCWs. This belief was significantly less concerned with citizens who needed hospitalization because of covid-19 infection, which might be due to their own possible need for ventilation.

Regarding the criterion of disconnecting the ventilator from a poor prognosis patient in favor of connecting it to a patient with a better prognosis, not agreement nor disagreement got the majority of votes among citizens and HCWs. However, HCWs agreed more on that. One of the jurisprudential challenges of connecting a ventilator to a patient is that disconnecting it is considered as murder from the perspective of Shiite jurists in Iran. Although this is not their opinion in severe shortage of ventilators, this matter is not well discussed and documented. The legal system in Iran is basically based on Shiite school of Islamic jurisprudence, so there is no legal protection for doctors from civil liability if they withdraw a ventilator to priorities the clinical need of another patient who could be saved. This challenge seems not to be unique to our country. For example, ventilator allocation guidelines in UK have not well addressed this problem(24). According to the general belief, disconnecting the device creates a more negative moral sense (25). However, in public health crises, the proper use of resources and avoiding waste of resources for patients who do not respond appropriately to the treatment should be considered. Therefore, it is necessary to create a public discourse on this criterion (25) and solve its jurisprudential challenge in dialogue with jurists. While many bioethicists and philosophers believe that there is not significant difference between withholding and withdrawing life-sustaining treatments from an ethical perspective (26).

The statement “Someone who needs the device sooner, has the right to use it even if he is less likely to survive than the next patient” is the sort of reverse statement of disconnecting the device in favor of the patient with a better prognosis. The opinion of majority of citizens’ and the HCWs was against this statement, although it still contained less than 50% of the opinions. These findings are close to Biddison et all study which showed that 43.7% of lay people and 59.5% of HCWs are against this criterion although their study was done in a no critical supply shortage time (7). Based on citizens’ concerns about incorrect diagnosis and early or inappropriate disconnection of the ventilator from patients, it seems necessary to take into account 1) criteria for excluding patients who have clearly weak prognosis and are unlikely to respond to ICU care based on good level of evidences, 2) reassessment of patient’s response to ICU care, and 3) explaining the policy of reassessment of patient for continuation of ventilation to patients and their family on admission(27).

The statement for priority based on lower age between two patients with the same prognosis, was based on the principle of the life cycle. In this study, a difference of 20 years between two patients was considered, and in the text of the statement, this difference of twenty years was raised both between the young and middle-aged patients and between the middle-aged and elderly patients. The results of this study showed that there is no meaningful agreement on the priority of a person who is 20 years younger. This finding may indicate that people do not accept the life cycle as a fair criterion, or that a 20-year age difference is not a good symbol for a life cycle and this amount of difference is not worth for prioritizing patients. Other surveys show that age is a very controversial criterion for allocating resources from people’s perspectives. In a study in Cyprus, the patient’s age (in favor of younger people) was an important criterion for health resource allocation(28), while public opinion polls from several European countries (Germany, the Netherlands, France, Italy, the United Kingdom, and Sweden) showed that people in all countries of the review were in disagreement with considering younger age as a basis of allocating health resources(29). A recent experiment showed that engaging participants in veil-of-ignorance reasoning changes their preferences toward saving younger patients(30). On the other hand, studies show that involving people in decision-making to allocate resources in an epidemic leads to their support for the life-cycle criterion(5, 25). In order to consider the life cycle in prioritization, it seems necessary to discuss the ethical basis of this criterion in public debate(2), and in case of acceptance, a consensus should be made on the valuable age difference.

The study had a number of limitations, including the data collection method which was electronic and non-random sampling through social media. This sampling method prevented the participants from being distributed by the education level in a ration of the general public, and people with higher education and those with more access to information technology were more likely to participate in this study. However, the analysis of the study results showed that people from all provinces of the country participated in this study and the distribution of samples of this study in different areas of technology development was not different from the distribution of the country’s general population in these areas.

## Conclusion

The results of this study showed that majority of people agreed upon the need to allocate resources to maximize health benefits in an emerging public health crisis through prioritizing higher prognosis patients. However, there is no agreement on reallocation of ventilators in the event of a resource shortage crisis. The criterion for periodic assessment of the status of patients and disconnecting ventilator device from a patient who does not benefit from ICU services requires its scientific and ethical basis to be brought in public discourse.

Although the panic from the shortage of resources in the pandemic of COVID-19 has decreased, the crisis is not over yet. In addition, this infectious epidemic will not be the last to occur and challenge the health care system of countries. So, in addition to look for more resource supply, efficient allocation of resources should be planned. It is necessary to involve the public in the discussions for this planning before the epidemic crisis. Therefore, in the event of a crisis and the need for health services, resource limitations would not lead to arbitrary allocation decisions, which reduces public confidence in health professionals and the health system.

## Data Availability

Anonymous data of this study is available upon request.

## Footnotes

1 https://app.epoll.pro/

## Notes

### Competing Interest Statement

The authors have declared no competing interest.

### Funding Statement

This paper is based on a self-funded study.

### Author Declarations

The study protocol was approved by the research ethics committee of Tehran University of Medical Sciences under license ID IR.TUMS.VCR.REC.1399.085.

